# Studies to develop a glucagon sensitivity test in humans: The GLUSENTIC study protocol

**DOI:** 10.1101/2022.11.05.22281981

**Authors:** Sasha A. S. Kjeldsen, Michael M. Richter, Nicole J. Jensen, Malin S. D. Nilsson, Niklas Heinz, Janus D. Nybing, Frederik H. Linden, Erik Høgh-Schmidt, Mikael P. Boesen, Sten Madsbad, Hendrik Vilstrup, Frank Vinholt Schiødt, Andreas Møller, Kirsten Nørgaard, Signe Schmidt, Elias B. Rashu, Lise L. Gluud, Steen B. Haugaard, Jens J. Holst, Jørgen Rungby, Nicolai J. Wewer Albrechtsen

**Affiliations:** Novo Nordisk Foundation Center for Protein Research, Faculty of Health and Medical Sciences, University of Copenhagen, Copenhagen, Denmark; Department of Clinical Biochemistry, Bispebjerg and Frederiksberg Hospital, Bispebjerg, Denmark; Department of Biomedical Sciences, Faculty of Health and Medical Sciences, University of Copenhagen, Copenhagen, Denmark; Department of Endocrinology, Bispebjerg University Hospital, Copenhagen, Denmark; Department of Radiology, Bispebjerg University Hospital, Copenhagen, Denmark; Department of Endocrinology, Hvidovre University Hospital, Hvidovre, Denmark; Department of Hepatology and Gastroenterology, Aarhus University Hospital, Aarhus, Denmark; Department of Gastroenterology, Bispebjerg University Hospital, Copenhagen, Denmark; Department of Gastroenterology and Gastrointestinal Surgery, Hvidovre University Hospital, Hvidovre, Denmark; Novo Nordisk Foundation Center for Basic Metabolic Research, Faculty of Health and Medical Sciences, University of Copenhagen, Copenhagen, Denmark; Institute of Clinical Medicine, Faculty of Health Science. University of Copenhagen; Steno Diabetes Center Copenhagen, Herlev, Denmark

**Keywords:** Non-alcoholic fatty liver disease, type 1 diabetes, Glucagon, Amino acids, Metabolism, The liver-alpha cell axis, Study Protocol

## Abstract

**Introduction:** A physiological feedback system exists between hepatocytes and the alpha cells termed the liver-alpha cell axis and signifies the role between amino acid-stimulated glucagon secretion and glucagon-stimulated amino acid catabolism. Several reports indicate that metabolic diseases such as non-alcoholic fatty liver disease (NAFLD) disrupts this feedback system, because of impaired glucagon receptor (GCGR) signaling (glucagon resistance). However, no experimental test exists to assess glucagon resistance in humans.

**Objective:** To develop and evaluate a test for measuring glucagon sensitivity towards amino acid and glucose metabolism in humans.

**Methods and analysis:** The study protocol is based on several pilot studies presented in this paper. The study will include 65 participants including 20 individuals with a BMI 18.6-25 kg/m^2^, 30 individuals with a BMI ≥25-40 kg/m^2^, and 15 individuals with type 1 diabetes with a BMI between 18.6-40 kg/m^2^. Participants will be grouped according to their percentage of hepatic steatosis measured by whole-liver magnetic resonance imaging (MRI). The primary outcome measure will be differences in a novel ‘glucagon sensitivity’ index between individuals with and without hepatic steatosis (<5.6 % vs ≥5.6 %) without diabetes. Secondary outcomes include between-group differences regarding the glucagon-alanine-index, incremental and decremental area under the curve (AUC) and association analyses between hepatic steatosis and glucagon sensitivity. This report describes the design of the cross-sectional study currently taking place at Copenhagen University hospital Bispebjerg and Frederiksberg.

**Results:** These data will be published in peer-reviewed scientific journals and presented at scientific conferences.

**Ethics and dissemination:** The study was approved by the scientific-ethical committee of the Capital region of Denmark (H-20023717) and registered with Danish Data protection Agency (P-2021-39) and ClinicalTrials.gov (NCT04907721). Written and oral consent will be obtained from all participants, and the study will adhere to the principles of the Declaration of Helsinki.

**Strengths and limitations of this study:**

- The glucagon sensitivity test is based on several pilot experiments
- Liver fat is based on whole-liver imaging and not region of interest (ROI)
- The glucagon sensitivity test may be limited to assess glucagon sensitivity towards amino acid catabolism and glucose production
- The glucagon sensitivity test does not use amino acid or glucose tracers which expands the generalizability of such test but also may impair its accuracy

## Introduction

Glucagon regulates amino acid turnover and amino acids may control glucagon secretion (1-3) in a feedback loop recently termed the liver-alpha cell axis (4). Pharmacological disruption of glucagon receptor (GCGR) signaling leads to increased plasma concentrations of amino acids (hyperaminoacidemia) and glucagon (hyperglucagonemia) in animals (5-12). This phenotype is also evident in rodents with hepatic steatosis (7) and, importantly, in humans with non-alcoholic fatty liver disease (NAFLD) (13, 14). Hyperglucagonemia and hyperaminoacidemia are believed result from a disrupted liver-alpha cell axis by which glucagon-stimulated amino acid catabolism is reduced and leads to hyperaminoacidemia, which, via known and unknown mechanisms, stimulates glucagon secretion from the alpha cells. NAFLD may impair glucagon’s effect on amino acid metabolism i.e., reduce glucagon sensitivity.

Our study protocol includes two experimental days, designed to evaluate the impact of both exogenous and endogenous glucagon on amino acid metabolism. Glucagon sensitivity will be evaluated using the glucagon-alanine index and a novel glucagon sensitivity index, described here. We will apply this test in patients with and without NAFLD and in individuals with type 1 diabetes. The latter group will serve as a model to investigate the dependency of varying insulin concentrations on the results of the test. The duration and dose of the amino acid infusion and glucagon injection included in our study protocol are based on previous human studies (15-17) and pilot studies presented here. The pilot studies were conducted in healthy individuals, where amino acid infusion rates, doses of glucagon infusions and sampling time have been evaluated. Furthermore, effects of single bolus injections of glucagon given during both fasted and non-fasted conditions are presented in this paper.

## Materials and methods

### Study approvals and ethical considerations

The study was approved by the scientific-ethical committee of the Capital region of Denmark (H-20023717) and registered with Danish Data protection Agency (P-2021-39) and ClinicalTrials.gov (NCT04907721). Written and oral consent is obtained from all participants, and the study will adhere to the principles of the Declaration of Helsinki.

### Pilot studies

The study protocol is based on several pilot studies investigating dose, time frames, and the administration of glucagon and amino acids individually or in combination. Nine pilot studies were conducted (named pilot studies 1-9).

In the first two pilot studies, a low dose of mixed amino acids (Vamin 14 g/l Electrolyte Free, catalog no. B05ABA01; Fresenius Kabi, Copenhagen, Denmark, 292 mg/kg BW/hour) was administered intravenously for 30 min in pilot study 1 and for 45 min in pilot study 2. A bolus injection of glucagon (1 mg) was administered 30 min after the amino acid infusion was stopped in pilot study 1, while a bolus injection of glucagon (0.5 mg) was given immediately after the amino acid infusion was stopped at time 45 min in pilot study 2. Two healthy individuals underwent pilot study 1 and 2 with one person completing both pilot studies. The purpose of pilot study 1 and 2 was to determine whether exogenous glucagon could accelerate amino acids disappearance, to evaluate dosing of glucagon and to identify a steady state plateau for plasma amino acids.

The same three healthy individuals completed pilot studies 3-6 (overview shown in Figure 3). In pilot studies 3-5, subjects received a high rate amino acid infusion of 370 mg/kg bw/hour similar to a previous report (18) from time 0-90 min. A bolus injection of glucagon (0.2 mg) was given at time 60 min (during the amino acid infusion) in pilot study 3 and at time 90 min (immediately after the amino acid infusion was stopped) in pilot study 4. In pilot study 6, amino acids were infused at a lower rate (331 mg/kg BW/hour) from 0-90 min equivalent to the middle of the two amino acid infusions investigated in pilot study 1 and 3. Three individuals completed the final pilot studies 7-9 in different order. Pilot study 7 consisted of a bolus injection of glucagon (0.2 mg) following an overnight fast, while in pilot study 8 participants ingested a breakfast two hours prior to a bolus injection of glucagon (0.2 mg). Pilot study 9 consisted of a saline following an overnight fast. No statistical analysis has been made of the pilot studies due to low sample size (n = 2-3).

### Pilot study results

Plasma amino acid levels were measured in pilot study 1 and 2 (Figure 1A-1B). When comparing the two pilot studies, it appeared that a bolus injection of glucagon accelerated the fall in plasma levels of amino acids following a state of high substrate availability (Figure 1C). Amino acid levels increased by 1.7 mmol/L to a peak value of 3.4 mmol/L, while endogenous glucagon levels increased by 12 pmol/L (from baseline to 16 pmol/L; data not shown). The increase in plasma levels of amino acids was comparable to those obtained in previous studies (19) following the ingestion of a protein-rich meal. This prompted us to explore the use of a higher amino acid infusion rate since between-group comparisons would become more apparent when evaluating larger responses. Plasma levels of glucose, insulin and C-peptide invariably increased in response to exogenous glucagon in pilot study 1 and 2 (Figure 2A-F), however, a larger increase was evident in pilot study 1 when plasma levels of amino acid levels were lowered (2.4 mmol/L vs 3.2 mmol/L in pilot study 2).

**Figure 1.**
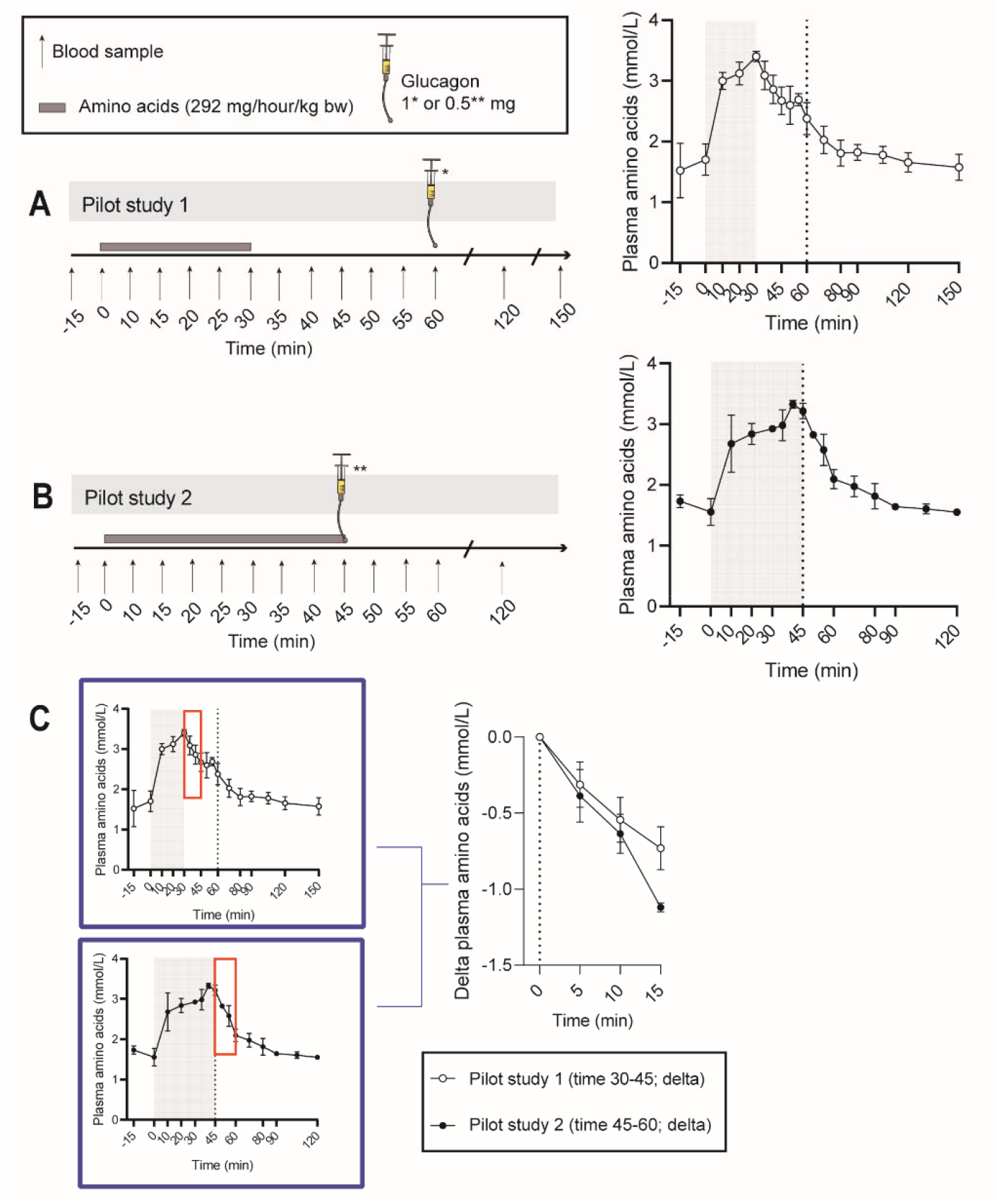
Exogenous glucagon (as indicated by vertical dotted line; 1 mg in pilot study 1 and 0.5 mg in pilot study 2; GlucaGen Hypokit Novo Nordisk) accelerates the decline in amino acid levels following an amino acid challenge (as indicated by the grey area; 292 mg/hour/kg bw; Vamin 14 g/L Electrolyte Free) in lean and healthy persons. Protocol for A)pilot study 1 and **B**) pilot study 2 with corresponding plasma amino acid levels. **C**) Plasma levels of amino acids decline more in response to exogenous glucagon following an amino acid challenge. Two healthy individuals completed pilot study 1 (BMI 22 ± 4 kg/m^2^, f/m 2/0) and pilot study 2 (BMI 21 ± 3 kg/m^2^, f/m 1/1) with one individual completing both pilot studies. Data are shown as mean ± SEM.

**Figure 2.**
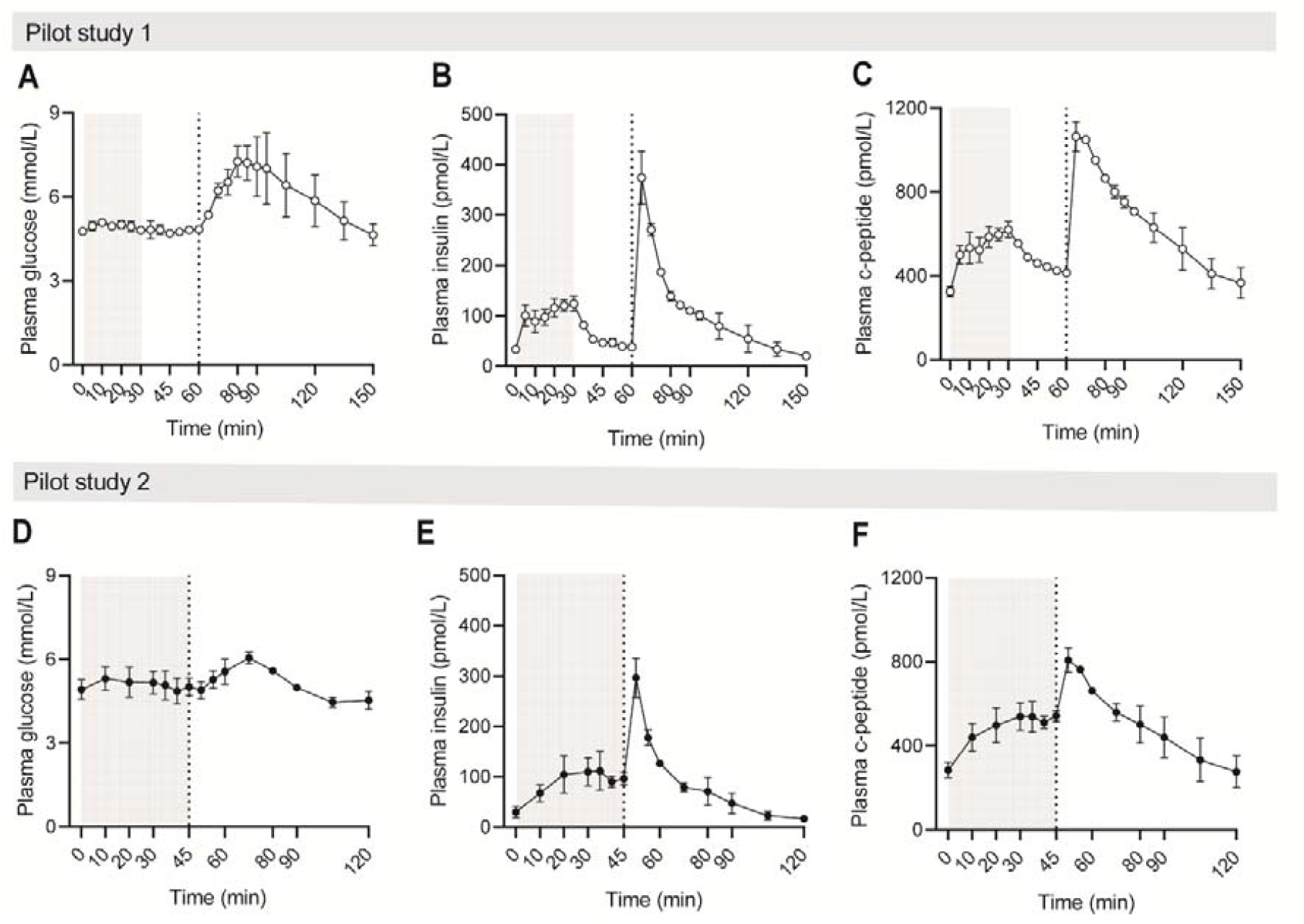
**A-F**) Plasma levels of glucose, insulin and c-peptide for pilot studies 1 and 2. A low amino acid infusion rate of 292 mg/hour/kg bw (as indicated by the grey area; Vamin 14 g/L Electrolyte Free) was infused for 30 min in pilot study 1 and 45 min in pilot study 2. As indicated by the vertical dotted line, a bolus injection of glucagon (GlucaGen Hypokit Novo Nordisk) was given 30 min after the amino acid infusion was stopped in pilot study 1 (1 mg) and immediately after the amino acid infusion was stopped in pilot study 2 (0.5 mg). Two healthy individuals completed pilot study 1 (BMI 22 ± 4 kg/m^2^, f/m 2/0) and pilot study 2 (BMI 21 ± 3 kg/m^2^, f/m 1/1) with one individual completing both pilot studies. Data are shown as mean ± SEM.

**Figure 3.**
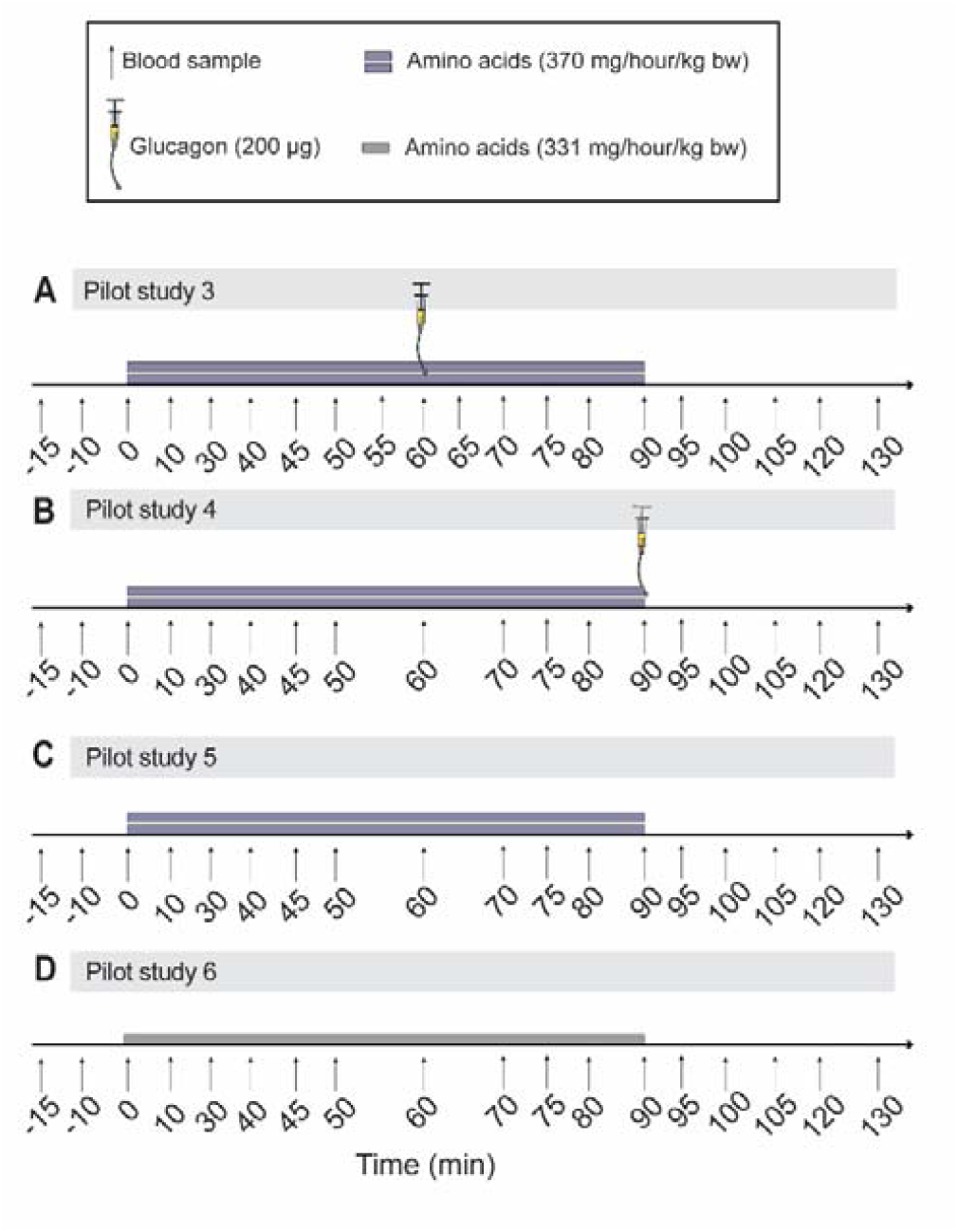
Pilot study protocols for pilot studies 3-9. The same three healthy participants (BMI 23 ± 2 kg /m2, f/m 1/2) completed pilot studies 3-6 (**A-D**). In pilot studies 3-5 (**A-C**), a high rate amino acid infusion (Vamin 14 g/L Electrolyte Free) of 370 mg/hour/kg bw was administered over 90 min, and in pilot study 6 (**D**) amino acids were infused at a rate of 331 mg/hour/kg bw. A bolus injection of glucagon (0.2 mg; Novo Nordisk GlucaGen Hypokit) was administered during the amino acid infusion at time 60 in pilot study 3 (**A**), while the bolus injection of glucagon was given immediately after the infusion was stopped in pilot study 4 (**B**).

In pilot studies 3-6, we further explored glucagon in combination with an amino acid infusion and increased the amino acid infusion rate to a rate previously administered in humans (18) and lowered the bolus infusion of glucagon to 0.2 mg (Figure 4A-P) due to mild nausea. Exogenous glucagon administered during (time 60 min; Figure 4A), but also immediately after (time 90 min; Figure 4E) the amino acid infusion (infused from time 0-90 min), did not alter plasma levels of amino acids when compared to the amino acid infusion alone (Figure 4I).

**Figure 4.**
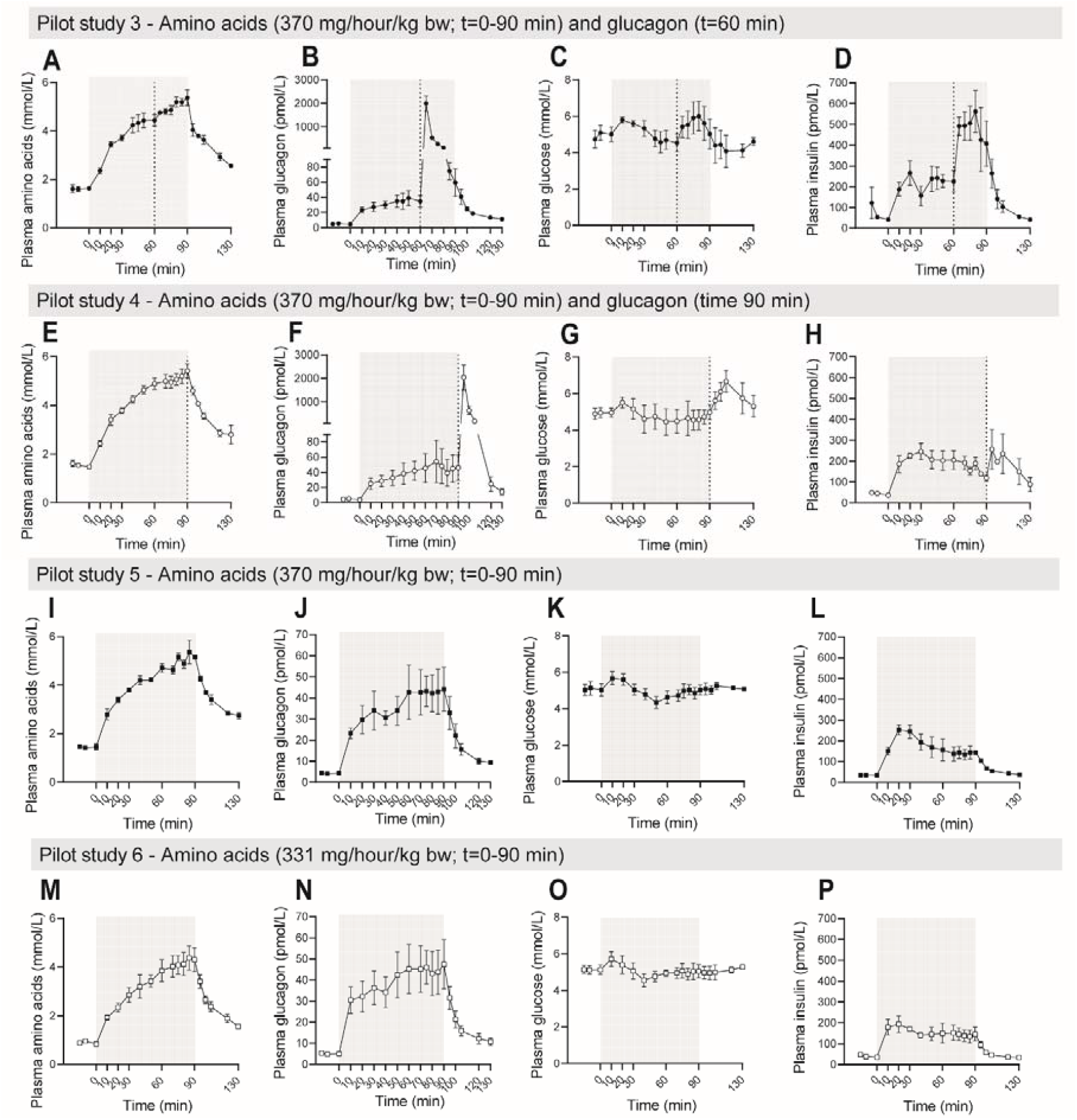
**A-P)** Plasma levels of amino acids, glucagon, glucose and insulin for pilot studies 3A-3D. In 3A, 3B and 3C (**A-L**) amino acids were infused at a rate of 370 mg/hour/kg bw over 90 min (as indicated by the grey area; Vamin 14 g/L Electrolyte Free) while a lower infusion rate of 331 mg/hour/kg bw was employed for 90 min in 3D (**M-P**). A bolus injection of glucagon (as indicated by the vertical dotted line; GlucaGen Hypokit Novo Nordisk) was given at time 60 min in pilot study 3A (**A-D**) and immediately after the amino acid infusion was stopped at time 90 min in pilot study 3B (**E-H**). Data are shown as mean ± SEM.

Next, we evaluated differences in metabolic parameters with the three amino acid infusion rates used in pilot studies 1, 2, 5 and 6 shown as baseline-subtracted (delta) values (Figure 5A-E). A low infusion rate of 292 mg/hour/kg bw was used in pilot studies 1 and 2, while a medium (331 mg/hour/kg bw) and high (370 mg/hour/kg bw) infusion rate was used in pilot studies 6 and 5. Plasma levels of amino acids increased dose-dependently, however, only a minor (if any) increase was observed when comparing the medium and high infusion rate (Figure 5A). Similarly, glucagon increased dose-dependently with the amino acid infusion rates, with little to no additional effect from the medium to high infusion rate (Figure 5C). A biphasic glucose response was observed for the medium and high amino acid infusion rates with an initial increase followed by a drop below baseline (Figure 5B). Finally, insulin and c-peptide increased dose-dependently with the amino acid infusion rates (Figure 5D-E).

**Figure 5.**
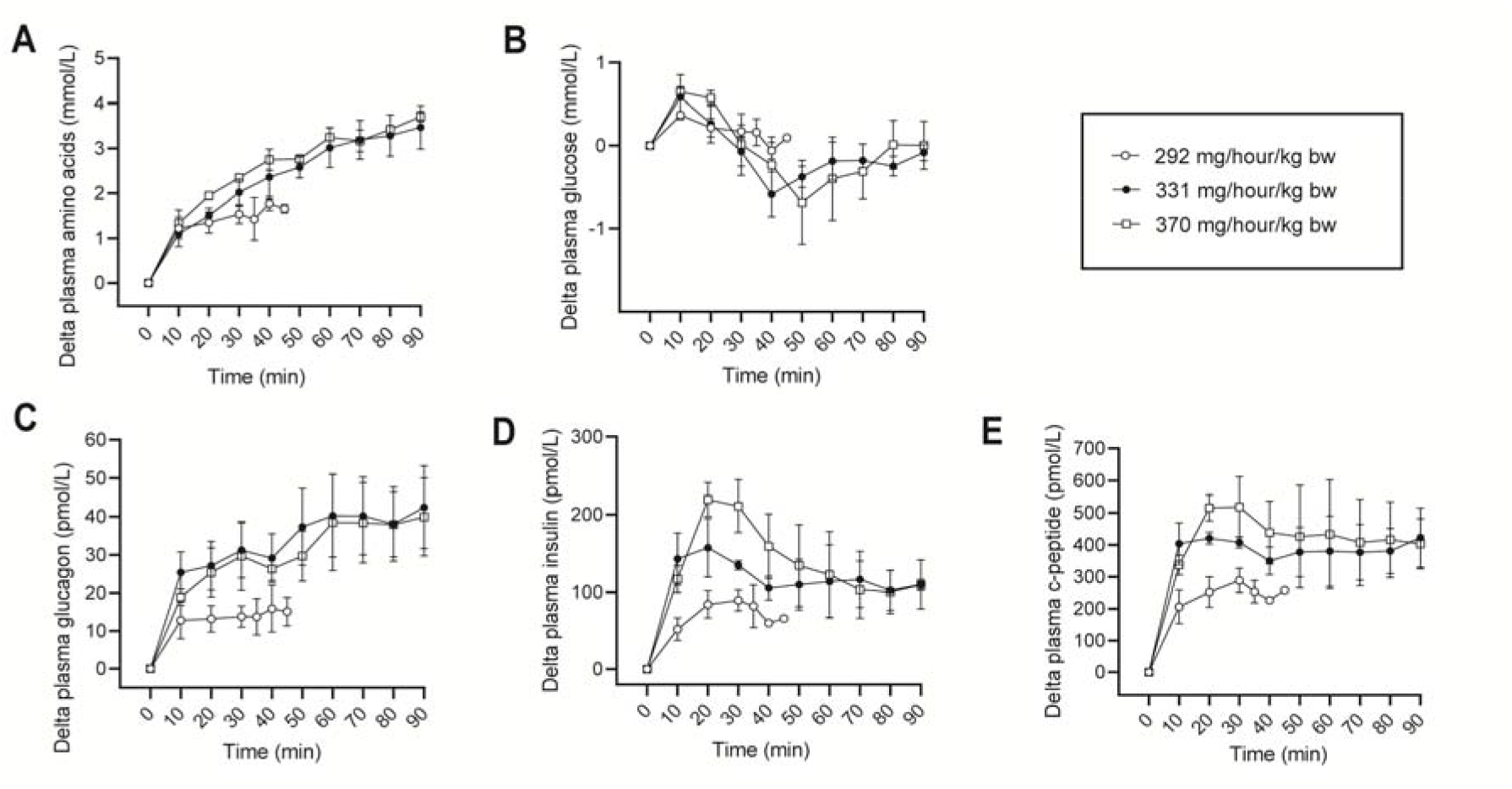
Comparing amino acid infusion rates from pilot studies 1, 2, 5 and 6. Plasma levels of **A**) amino acids, **B**) glucose, **C**) glucagon, **D**) insulin and **E**) c-peptide when comparing different amino acid infusion rates for pilot studies 1 and 2 (combined, n=3; 292 mg/hour/kg bw), pilot study 5 (n=3; 370 mg/hour/kg bw) and pilot study 6 (n=3; 331 mg/hour/kg bw). Amino acids (Vamin 14 g/L Electrolyte Free) were infused for 30-45 min in pilot studies 1 and 2, and for 90 min in pilot studies 5 and 6. Data are shown as mean ± SEM.

The same three individuals completed (in different order) pilot studies 7-9, consisting of a bolus injection of saline or glucagon after an overnight fast or glucagon two hours after a breakfast meal. In pilot study 9 (breakfast + glucagon), subjects are considered to be in the fed state as indicated by increased plasma levels of insulin at baseline (248 pmol/L following breakfast vs 45 pmol/L after an overnight fast). Plasma levels of amino acids fell following exogenous glucagon independently of the fed and fasted state. Plasma glucose levels increased following exogenous glucagon (Figure 6C-D); however, this increase was almost abolished following the breakfast meal, presumably because of the increased plasma insulin levels at baseline (Figure 6E-F). Plasma levels of urea did not fall to the same extent following exogenous glucagon (Figure 6G).

**Figure 6.**
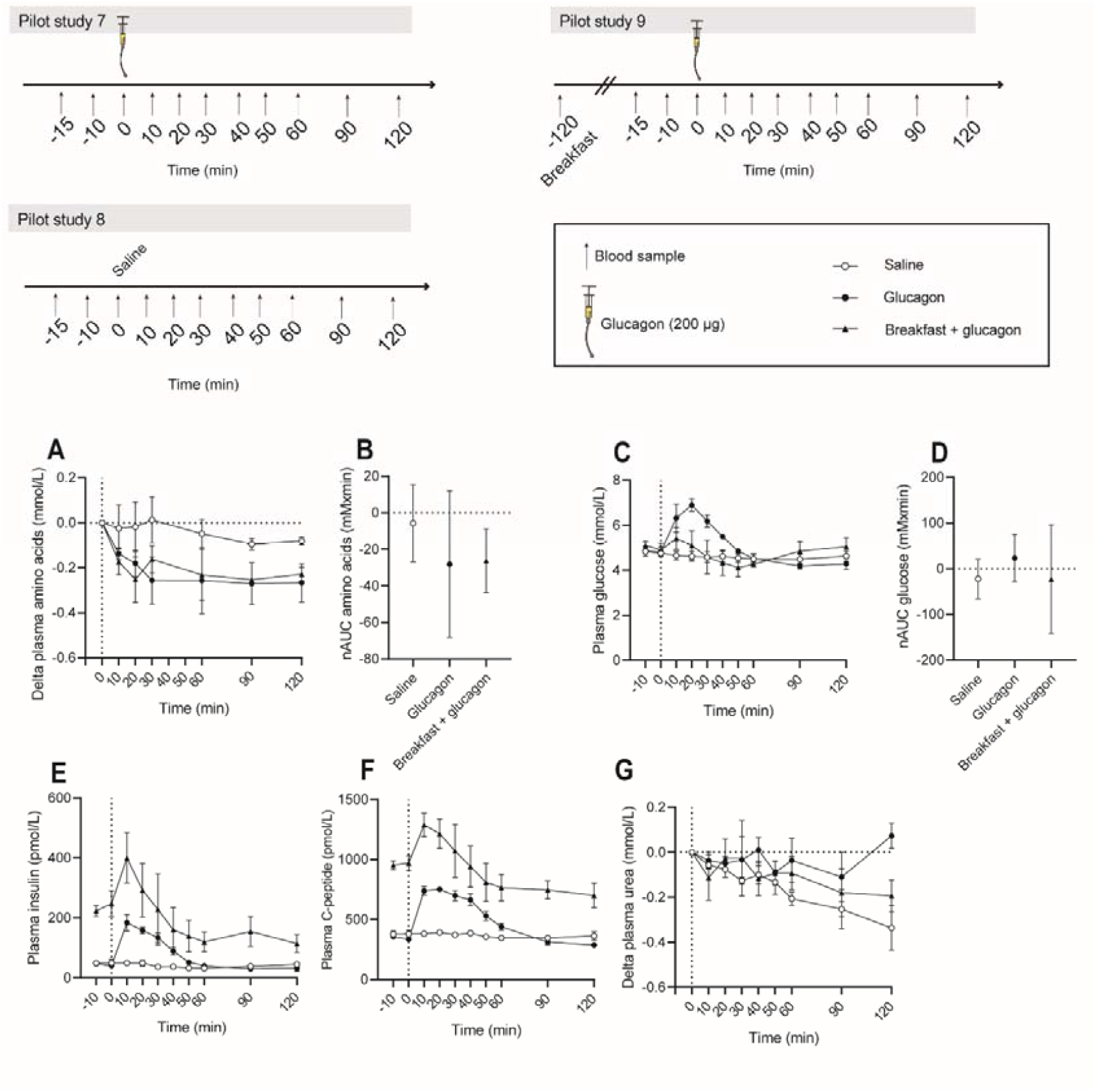
Plasma levels of **A-B**) amino acids, **C-D**) glucose, **E**) insulin, **F**) c-peptide and **G**) urea for pilot studies 7-9, completed by the same three healthy individuals (BMI 24 ± 1 kg /m2, f/m 3/0) on three separate days. A bolus injection of glucagon (GlucaGen Hypokit Novo Nordisk; as indicated by the vertical dotted line) was administered at time 0 after an overnight fast (black circles) or following a breakfast (80-100 g of oats, 220 ml skimmed milk and 30 g raisins) 2 hours prior (black triangles). On a third day, individuals received saline (open circles) at time 0 min after an overnight fast. Data are shown as mean ± SEM.

### Pilot study discussion

Plasma levels of amino acids decreased following a bolus injection of glucagon compared to saline (Figure 1).

Bolus injection of 0.2 mg glucagon caused a supra-physiological plasma glucagon level of 2000 pmol/L (Figure 4B and 4F) and maximum responses to glucagon are most likely reached at exogenous glucagon doses of 0.2 mg or lower.

Plasma levels of glucose increased more in response to glucagon when plasma levels of amino acids were low (43-50%; in pilot study 1 and 7) compared to high (21-33%; in pilot study 2, 3 and 4), presumably due to the amino acid-stimulated increase in insulin levels and subsequent insulin-stimulated tissue uptake of glucose. An increased glucagon dose did not change this since pilot study 2 (0.5 mg glucagon) elicited a 21% increase in glucose, while pilot study 4 (0.2 mg glucagon) elicited a 43% increase in glucose.

When evaluating changes in plasma amino acid levels with exogenous glucagon, amino acids only decreased when individuals had *not* been subjected to a very high amino acid infusion rate. Pilot study 3 and 4 show that exogenous glucagon had no effect on amino acid clearance both during and following a high amino acid infusion rate, while a reduction in plasma amino acid levels was evident in pilot study 2, 7 and 9 following a low amino acid infusion rate or no amino acid infusion at all. This difference may be the result of saturation of the GCGR, i.e., maximal stimulation of ureagenesis by the amino acid-stimulated secretion of endogenous glucagon rendering additional exogenous glucagon incapable of further increasing ureagenesis. Based on these observations, we decided that the GLUSENTIC protocol should consist of two experimental days with the aim of examining the effect of both exogenous and endogenous (amino acid-stimulated) glucagon in individuals with and without NAFLD. In prospective studies it may turn out that glucagon sensitivity may be adequately evaluated based on a single glucagon injection.

## The GLUSENTIC study protocol

Based on the pilot studies described above, the final study protocol consists of two experimental study days to take place within two weeks of each other. Participants will perform both study days in the morning following an overnight fast from 9:30 PM the night before (participants with type 1 diabetes are exempted from this) and will remain fasted throughout the study day but with access to water. Each study day will have a duration of approximately 4 hours. Participants will be asked to abstain from any strenuous physical activity and alcohol consumption for 48 hours prior to both study days. On both study days, an intravenous catheter will be inserted into an antecubital vein of both arms; one for infusion and one for blood sampling.

### Experimental day 1

On the first study day, participants will be scanned by magnetic resonance imaging (MRI). Liver fat, subcutaneous fat and visceral fat will be assessed with a commercially available version of a 6-point mDIXON sequence package done in 2-3 breath-holds covering the entire abdominal cavity. Participants will be scanned using a SENSE body array receive coil. Liver fat assessments will be based on a fully automated fat fraction quantification of the entire liver volume, using a dedicated software that calculates MRI biomarkers (liverhealth intellispace 11, Philips Healthcare Nederlands)(20). Visceral fat will be assessed by calculating the volume of visceral fat in the entire abdominal cavity, based on semi-automated segmentation approach (MANGO software)(21). In addition, anatomical images of the entire abdomen will be acquired using a 3D isotropic T1 weighted MRI sequence. The acquisition time of the MRI scan will be approximately 30 minutes in total (including preparation time). Only individuals with no MRI contraindications (e.g. metallic foreign bodies, MRI unsafe implants, claustrophobia) will be included. Subjects will be scanned in a 1.5 Tesla Ambition MRI scanner (Philips Healthcare, The Netherlands). Body composition will be evaluated using a validated bioelectrical impedance analysis (BIA), and waist and hip circumferences will be measured. Participants will then rest in a supine position. Following the placement of the two venous catheters, baseline blood samples will be taken at time points −10 and 0 min, immediately followed by a bolus injection of glucagon (0.2 mg; Novo Nordisk Glucagen Hypokit) at time 0 min after which blood will be sampled at time points 10, 20, 30, 60, 90 and 120 min (Figure 2).

### Experimental day 2

On the second study day, completed within 14 days of the first experimental day, participants will rest in a supine position and receive an intravenous infusion of amino acids (Vamin 14 g/l Electrolyte Free, catalog no. B05ABA01; Fresenius Kabi, Copenhagen, Denmark) from 0-45 min at an infusion rate of 3.885 ml/kg BW/hour corresponding to 331 mg/kg BW/hour. Blood samples will be obtained from −10 to 180 min (Figure 7)

**Figure 7.**
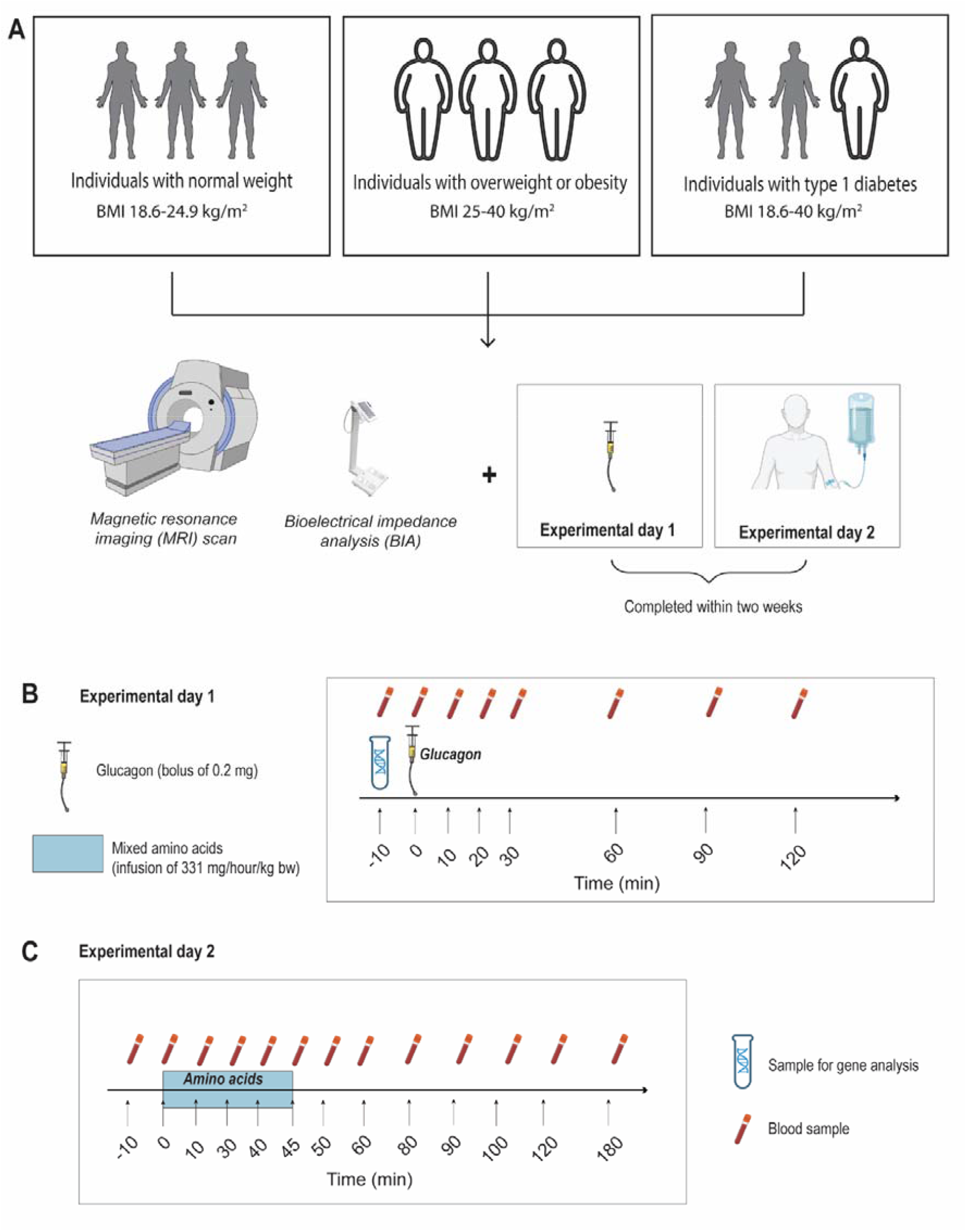
The GLUSENTIC study protocol. Final study protocol based on pilot studies 1-9. Three groups of individuals will complete two experimental study days. On the first study day, a bolus infusion of glucagon (0.2 mg; Novo Nordisk GlucaGen Hypokit) will be administered at time 0 min; while on the second study day, an amino acid mixture (331 mg/hour/kg bw; Vamin 14 g/L Electrolyte Free) will be infused for 45 min from time 0-45 min. Participants will be scanned by magnetic resonance imaging (MRI) and bioelectrical impedance analysis (BIA) and a blood sample for gene analysis will be obtained on the first experimental day.

### Participants

Three groups of subjects will be included in the GLUSENTIC study: Individuals with normal weight (BMI 18.6-25 kg/m^2^), individuals with overweight and obesity (BMI ≥ 25 kg/m^2^), and individuals with type 1 diabetes (BMI 18.6-40 kg/m^2^). Participant screening will include medical history, physical examination, and standard baseline laboratory tests. These three groups will – following study completion -be segregated into four groups based on hepatic fat content measured by MRI. Participants who drop out of the study before completion may be replaced by a new participant. Inclusion and exclusion criteria for lean individuals and individuals with overweight and obesity are summarized in table 1, while inclusion and exclusion criteria for individuals with type 1 diabetes are summarized in table 2.

**Table 1:**
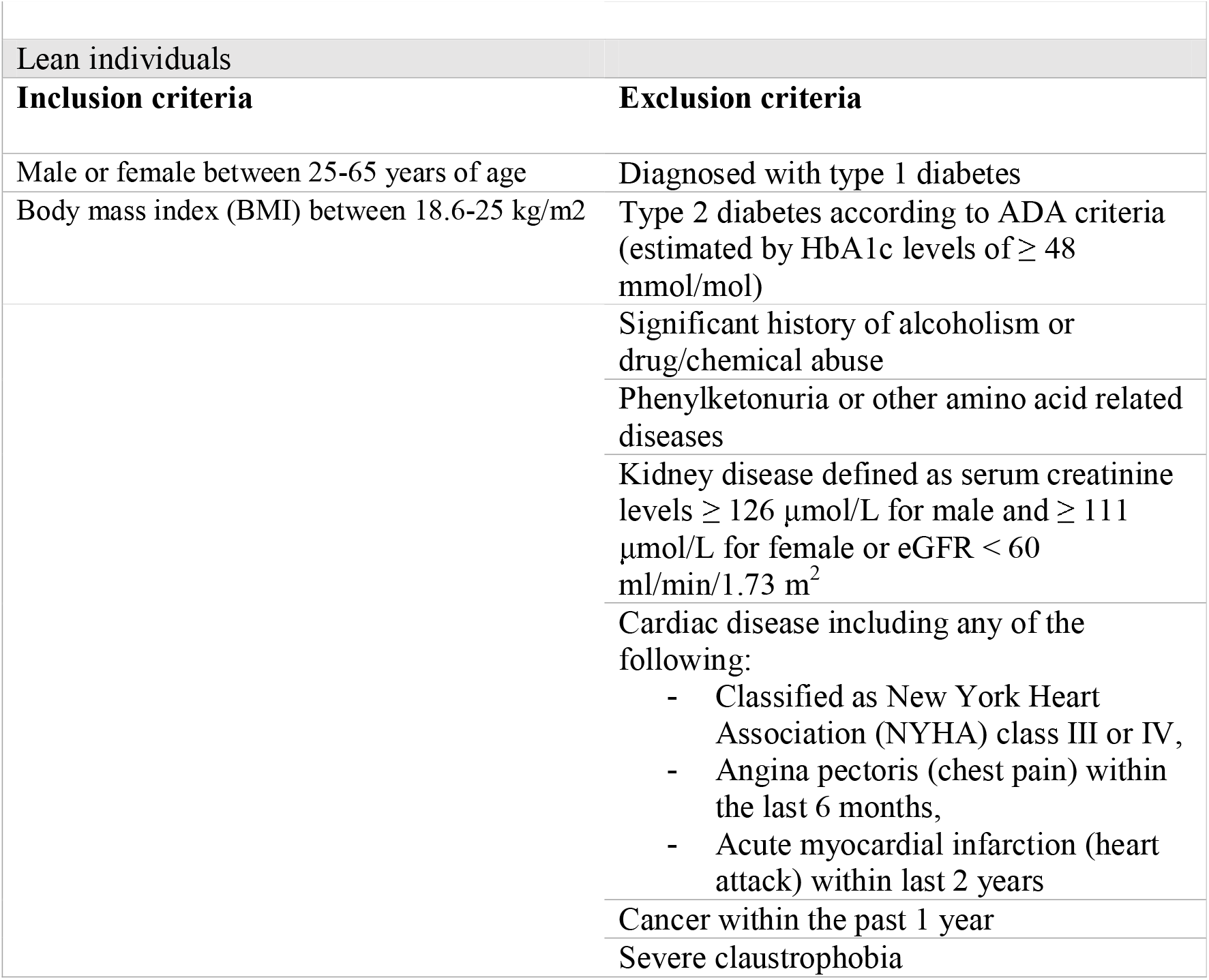
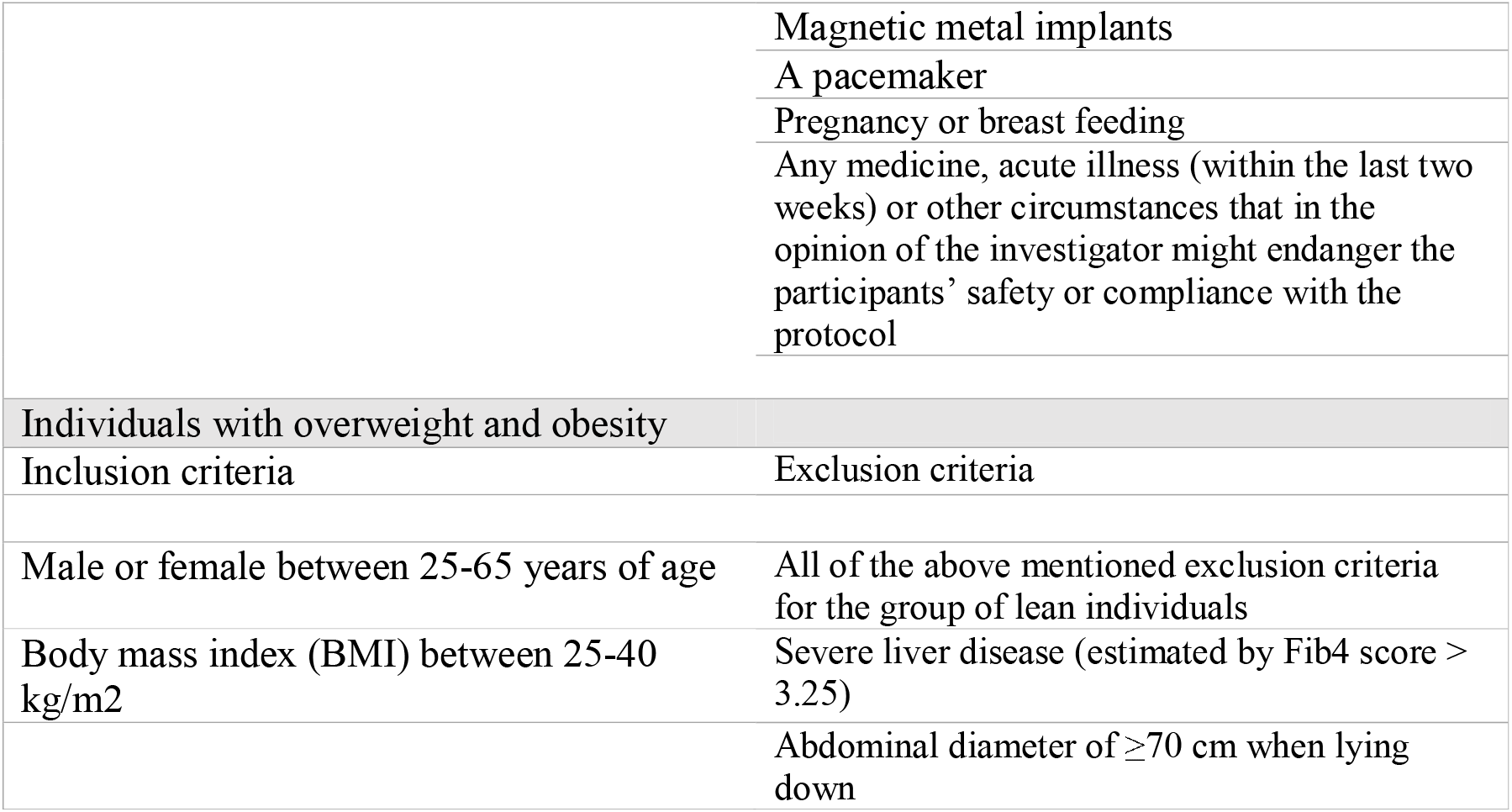
Inclusion and exclusion criteria for individuals with normal weight and individuals with overweight and obesity.

**Table 2:**
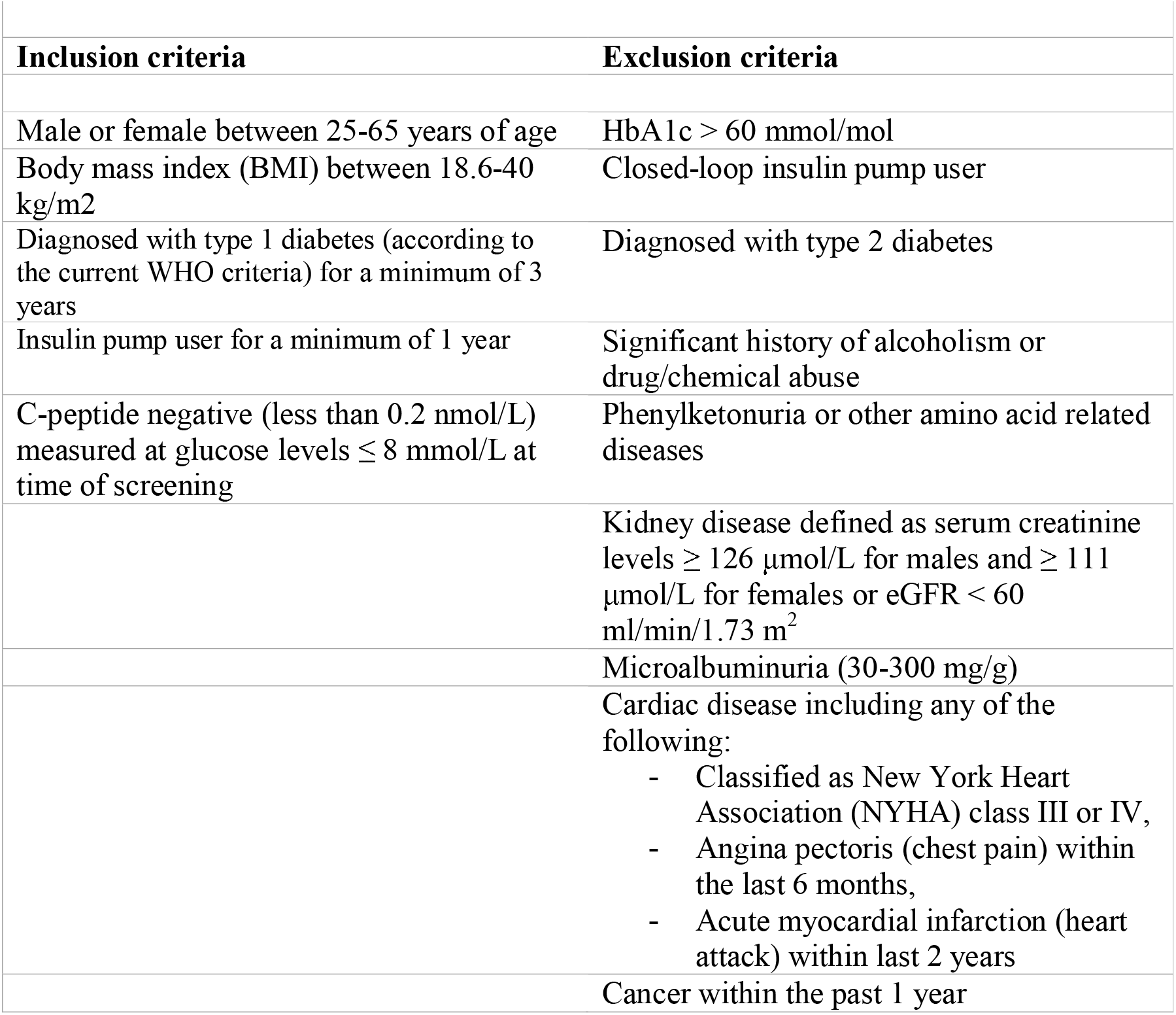
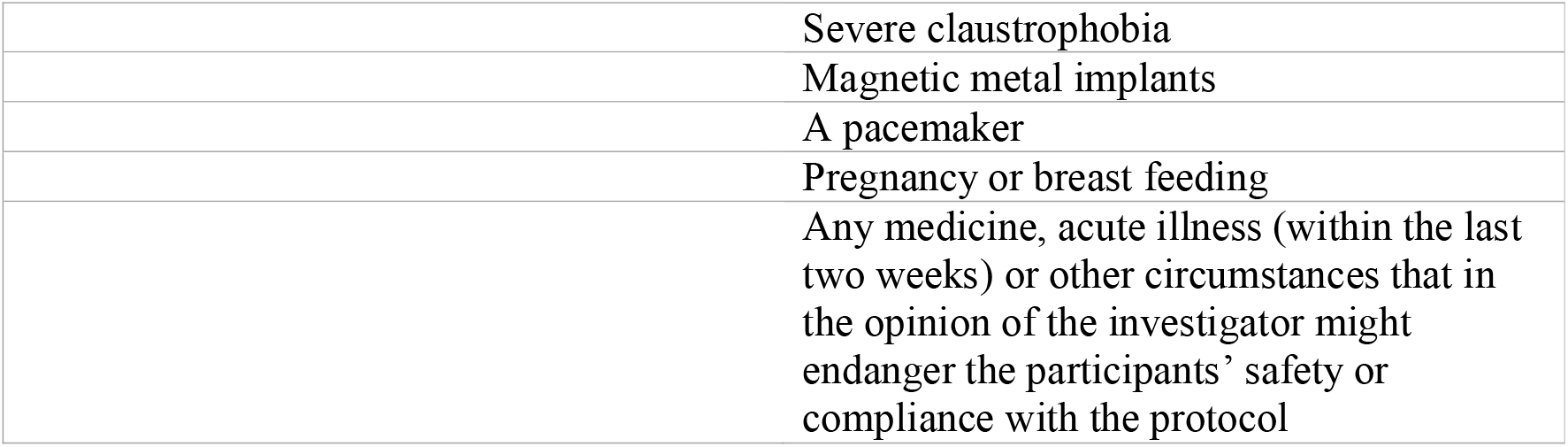
Inclusion and exclusion criteria for individuals with type 1 diabetes.

|  |  |
| --- | --- |
| Inclusion criteria | Exclusion criteria |
| Male or female between 25-65 years of age | HbA1c > 60 mmol/mol |
| Body mass index (BMI) between 18.6-40 kg/m<sup>2</sup> | Closed-loop insulin pump user |
| Diagnosed with type 1 diabetes (according to the current WHO criteria) for a minimum of 3 years | Diagnosed with type 2 diabetes |
| Insulin pump user for a minimum of 1 year | Significant history of alcoholism or drug/chemical abuse |
| C-peptide negative (less than 0.2 nmol/L) measured at glucose levels $\leq 8$ mmol/L at time of screening | Phenylketonuria or other amino acid related diseases |
| | Kidney disease defined as serum creatinine levels $\geq 126$ $\mu\text{mol/L}$ for males and $\geq 111$ $\mu\text{mol/L}$ for females or eGFR < 60 ml/min/1.73 m<sup>2</sup> |
|  | Microalbuminuria (30-300 mg/g) |
|  | Cardiac disease including any of the following:<br>- Classified as New York Heart Association (NYHA) class III or IV,<br>- Angina pectoris (chest pain) within the last 6 months,<br>- Acute myocardial infarction (heart attack) within last 2 years |
|  | Cancer within the past 1 year |
|  | Severe claustrophobia |
|  | Magnetic metal implants |
|  | A pacemaker |
|  | Pregnancy or breast feeding |
|  | Any medicine, acute illness (within the last two weeks) or other circumstances that in the opinion of the investigator might endanger the participants' safety or compliance with the protocol |

### Insulin Dosing for Individuals with Type 1 Diabetes

Participants with type 1 diabetes will follow an identical protocol but with extra additions/precautions. Participants with type 1 diabetes will be asked to set an alarm for 3AM on the morning of both experimental days and measure their blood glucose. If blood glucose levels are less than 4.5 mmol/L, the participant should reduce their insulin infusion rate or consume dextrose. If blood glucose levels are more than 10 mmol/L, the participant should correct with an insulin bolus. If blood glucose levels are less than 3 mmol/L during the night or morning of the experimental day, the subject should receive dextrose, and the trial should be rescheduled for another day. Study participants will continue their basal infusion rate of insulin from their insulin pump during the two experimental days. On the day of the magnetic resonance imaging scan, the participant must remove the insulin pump prior to the scan. When the scan is complete, the participant may re-attach the insulin pump. The participant will then be asked to measure blood glucose and if blood glucose is more than 6 mmol/L, the participant should correct with a bolus infusion of insulin corresponding to the amount of time the participant has been without the insulin pump multiplied by the participant’s basal infusion rate of insulin.

## Sample size calculation

Based on pilot data reported here and on a previous study of individuals with NAFLD (22) we estimated a sample size of 15 in each group enabling difference of 10% by an unpaired test, a power of 90%, an alpha value of 5% and 20% variation in the measured outcome.

## Study outcomes

### Primary outcome measure

Our main outcome measure will be differences in the ‘glucagon sensitivity index’ presented in the formula below:

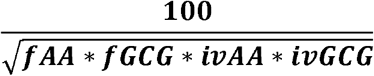

*fAA = Plasma levels of amino acids after an overnight fast (mean value at time −10 and 0)*.

*fGCG = Plasma levels of glucagon after an overnight fast (mean value at time −10 and 0)*.

*ivAA = Plasma levels of amino acids during the amino acid infusion (mean value at time 40 and 45)*.

*ivGCG = Plasma levels of glucagon during the amino acid infusion (mean value at time 40 and 45)*.

The glucagon sensitivity index is conceptually based on the Matsuda/composite index (23, 24) used to evaluate insulin sensitivity. We anticipate this index to be a novel marker for glucagon sensitivity and hence proper reference to this paper should be given when employed.

### Secondary outcomes

Differences in the glucagon-alanine index (un-paired t-test) measured after an overnight fast between individuals with (>5.6%) and without (<5.6%) hepatic steatosis who are not diagnosed with diabetes.

Differences in total amino acid levels after the bolus injection of glucagon calculated as the decremental AUC (time=0-120min). Comparisons between otherwise healthy individuals with and without hepatic steatosis will be made (unpaired t-tests), and comparisons between individuals with type 1 diabetes with and without hepatic steatosis will be made (unpaired t-tests).

Differences in plasma glucose levels after the bolus infusion of glucagon calculated as the incremental AUC (time=0-120min). Comparisons between otherwise healthy individuals with and without hepatic steatosis will be made (unpaired t-tests), and comparisons between individuals with type 1 diabetes with and without hepatic steatosis will be made (unpaired t-tests).

Differences in amino acid levels calculated as incremental or total AUC during the amino acid infusion (time=0-45min) (reflecting amino acid tolerance). Comparisons between otherwise healthy individuals with and without hepatic steatosis will be made (unpaired t-tests), and comparisons between individuals with type 1 diabetes with and without hepatic steatosis will be made (unpaired t-tests).

Differences in amino acid levels calculated as the decremental AUC after the amino acid infusion, reflecting amino acid clearance (time=45-180min). Comparisons between otherwise healthy individuals with and without hepatic steatosis will be made (unpaired t-tests), and comparisons between individuals with type 1 diabetes with and without hepatic steatosis will be made (unpaired t-tests).

Comparisons between non-diabetic individuals with and without hepatic steatosis will be made (unpaired t-tests), and comparisons between individuals with type 1 diabetes with and without hepatic steatosis will be made (unpaired t-tests).

Multiple linear regression between hepatic steatosis (% liver fat) and ‘glucagon sensitivity’ after adjusting for body weight and age. Regression analysis will be performed on data from otherwise healthy individuals with and without hepatic steatosis, and on data from individuals with type 1 diabetes with and without hepatic steatosis.

Differences in the following formula:

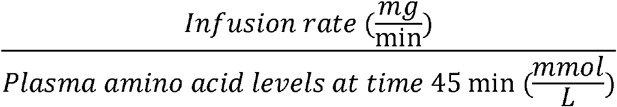

Comparisons between non-diabetic individuals with and without hepatic steatosis will be made (unpaired t-tests), and comparisons between individuals with type 1 diabetes with and without hepatic steatosis will be made (unpaired t-tests).

Rate of decay (time=45-180min): Delta values (baseline-subtracted values) for amino acids on a logarithmic scale transformed back to a relative change. Comparisons between non-diabetic individuals with and without hepatic steatosis will be made (unpaired t-test), and comparisons between individuals with type 1 diabetes with and without hepatic steatosis will be made (unpaired t-tests).

### Statistics

Area under the curve (AUC) will be calculated using the trapezoidal rule. Unpaired t-tests with post hoc correction for multiple testing will be used to compare fasting concentrations and AUCs. A mixed-effect model will be applied to test the effect of treatment (glucagon bolus or amino acid challenge) and time on the measured variables. Regression analysis between hepatic steatosis and glucagon sensitivity. Calculations and illustrations will be made using GraphPad Prism for Windows (GrapPad Software, La Jolla, CA) and R. Data will be shown as mean ± SEM if not otherwise indicated.

### Laboratory and liver fat analyses

Blood from the experimental day will be kept on ice and centrifuged according to guidelines. Plasma will be aliquoted and stored at −80°C until analysis and the sample containers/tubes will be labelled according to GDPR law. Glucagon will be measured using a two-site enzyme-linked immunoassay (cat. no. 10-1281-01; Mercodia, Upsala, Sweden) specific for intact C- and N-termini of the molecule according to the manufacturer’s protocol, and by an in-house radioimmunoassay (RIA) using antiserum (no. 4305), monoiodinated 125I-labeled glucagon, specific for the C-terminus of glucagon. Plasma concentrations of total L-amino acids will be measured using an enzymatic assay (catalog no. ab65347; Abcam, Cambridge, UK) according to the manufacturer’s protocol as well as an in-house assay measuring alpha-amino nitrogen. Plasma concentrations of insulin, C-peptide, glucose, and urea will be quantified with Cobas automated analyzer platform. Metabolomics (fractionated amino acids, lipid-species) will be measured by a mass-spectrometry based approach (25). TaqMan assays (an assay of specific predefined genes as opposed to exome sequencing) to identify single genetic variants, known from previous studies to affect the risk of liver disease: rs738409 in the gene *PNPLA3*; rs72613567 in the gene *HSD17B13*; rs641738 in the gene *MBOAT7*; rs1260326 in the gene *GCKR*, rs4841132 in the gene *PPP1R3B*, rs361525 in the gene *TNF*; and rs58542926 in the gene *TM6SF2* (26).

To obtain a valid measurement of liver fat we use the 6 point DIXON MRI PDFF methods enabling measurements of the whole liver fat content by use of the dedicated Liver analysis program Liverhealth intellispace 11, Philips Healthcare Nederlands(20), reducing the variation of liver fat estimation using the previously widely used “region of interest” analysis that is prone to inconsistent estimations (27, 28)

## Ethics and Dissemination

The study is approved by the scientific-ethical committee of the Capital region of Denmark (H-20023717) and registered with Danish Data protection Agency (P-2021-39) and ClinicalTrials.gov (NCT04907721). Written and oral consent will be obtained from all participants, and the study will adhere to the principles of the Declaration of Helsinki. The study will be conducted according to the experimental protocol.

In this study, we will investigate the hepatic response to an amino acid challenge and a bolus infusion of glucagon on two separate study days in individuals with and without NAFLD. Increased concentrations of glucagon (hyperglucagonemia), as seen in some individuals with type 2 diabetes, contributes to hyperglycemia. A test to evaluate glucagon sensitivity could be important to predict the development of type 2 diabetes. Furthermore, insulin sensitivity measurements have historically had significant impact on our way of interpreting metabolic dysfunction and diabetes, and a test for evaluating glucagon sensitivity may similarly prove to be an important tool. Based on the literature, we find that developing and evaluating a test for measuring hepatic glucagon sensitivity will be important for the understanding of glucagon biology and liver disease.

Discomfort and risk of complications are expected to be minimal. The study will not benefit the individual participant; however, the results could prove beneficial for future patients suspected of liver disease. Participants can choose whether they wish to receive information on chance findings. Should a participant be a carrier of one or more of the genetic variants measured in the present study, the concerned individual will be offered an informative interview on the test results and a specialist will take part in the initial conversation for counsel. Information on the findings including personal and family consequences will be discussed. The genetic mechanisms behind the disease will be explained and prognosis and treatment options will be discussed. Generally, the patient agrees that he or she informs his or her family members that a hereditary illness has been found in the family and that the health service offers an investigation to determine if the relatives may have inherited this disease as well. Counseling is offered individually or to the family.

Scientific publications are expected, and both positive, negative, and inconclusive results will be published. The manuscripts will be published in international medical journals on liver physiology, endocrinology, and metabolism. Data will be presented at national and international scientific conferences

## Data Availability

Data are avaliable upon request

## Data Availability Statement

Data are available through request to the corresponding author.

## Authors’ contributions

SASK, MMR, NJS, SM, HV, LLG, SBH, JJH, JR and NJWA conceived the idea of a glucagon sensitivity test and outlined the pilot studies and the final study protocol. MSD, NH, NDN, FHL, ERHS, MPB, FVS, KN, Sm, AM, EBR contributed with intellectual inputs to the protocol, assisted with patient recruitment, performance of experimental study days or performed imaging of the liver including data analyses. SASK, MMR and NJWA wrote the first draft of the manuscript. All other authors revised the manuscript and approved the final version.

## Acknowledgement

We appreciate Christine Rasmussen (Department of Clinical Biochemistry, Bispebjerg and Frederiksberg Hospital) for assistance with the study days and biochemical measurements. We thank our study participants for participating in the pilot studies.

